# Wastewater-informed agent-based modelling of hepatitis E transmission dynamics

**DOI:** 10.64898/2026.02.14.26346311

**Authors:** Karina Wallrafen-Sam, Jasmin Javanmardi, Nina Schmid, Mathias Schemmerer, Jürgen J. Wenzel, Andreas Wieser, Jan Hasenauer

**Affiliations:** Life & Medical Sciences Institute (LIMES), University of Bonn, Bonn, Germany; Bonn Center for Mathematical Life Sciences, University of Bonn, Bonn, Germany; Institute of Infectious Diseases and Tropical Medicine, LMU University Hospital Munich, Munich, Germany; National Consultant Laboratory for HAV and HEV, Institute of Clinical Microbiology and Hygiene, University Medical Center Regensburg, Regensburg, Germany; German Centre for Infection Research (DZIF), Partner Site Munich, Munich, Germany; Max von Pettenkofer Institute, Faculty of Medicine, LMU Munich, Munich, Germany; Immunology, Infection and Pandemic Research (IIP), Fraunhofer Institute for Translational Medicine and Pharmacology (ITMP), Munich, Germany

**Keywords:** Wastewater-based epidemiology, Infectious disease surveillance, Hepatitis E virus, Agent-based modelling, Epidemiological modelling, Bayesian calibration

## Abstract

Hepatitis E virus (HEV) is considered a predominantly foodborne pathogen in developed settings. During COVID-19 lockdown periods, however, HEV concentrations in wastewater at a treatment plant in Munich, Germany decreased, suggesting that pandemic-related behaviour changes inadvertently influenced transmission. In contrast, reported cases and wastewater data from a smaller catchment showed no comparable decline. To assess whether the observed reduction is compatible with a near-exclusively foodborne infection and to reconcile the contrasting signals across surveillance modalities, we developed a stochastic, individual-level model of HEV transmission, shedding, and ascertainment in Munich. Using Approximate Bayesian Computation, we calibrated this model to wastewater and case data from 2020–2023, first separately and then jointly. Posterior parameter estimates indicated a substantial decline in transmission during lockdowns to about 35–40% of the non-lockdown level, with the 95% credible interval entirely below 1 (no change). Joint inference suggested that possible modest lockdown-associated increases in diagnosis probabilities and higher measurement variability in the smaller catchment masked this effect in clinical and small-scale wastewater data, respectively. These findings demonstrate how wastewater-based surveillance, used alongside reported cases, can enable more robust parameter inference for models of under-reported pathogens like HEV, thereby supporting informed public health risk assessments.

## 1 Introduction

Wastewater-based surveillance (WBS) is emerging as a powerful tool for infectious disease surveillance that can offer population-level insights complementing those from traditional case-based reporting [1, 2]. WBS captures viral material shed by infected individuals regardless of their symptom or diagnosis status, making it less subject to under-reporting bias. This feature makes WBS particularly suitable for surveillance of hepatitis E virus (HEV), which can cause acute inflammatory hepatic infections in humans but is often clinically silent: Most HEV infections are mild or asymptomatic and thus remain undetected by clinical surveillance [3, 4]. In Germany, comparisons of seroprevalence data [5] to case reports [6] indicate that well over 95% of HEV infections are never reported. Nevertheless, HEV poses a growing public health concern due to recent reported case increases and the discovery of chronic hepatitis in immunosuppressed individuals [5, 7]. WBS thus provides a uniquely valuable means to assess HEV transmission dynamics at scale.

In developed countries where HEV genotype 3 predominates, transmission is understood to be mainly foodborne, especially through raw or undercooked pork products [3]. Foodborne infections are typically expected to be less sensitive to lockdown restrictions than infections spread via interpersonal contact. However, this assumption has recently been challenged by observations made during the COVID-19 pandemic: Analyses of wastewater data collected in Munich from 2020 to 2023 provided some evidence of reduced HEV concentrations during periods of strict lockdown, when social contacts were limited and restaurant closures altered food consumption behaviours [8]. Specifically, a statistically significant negative correlation was found between the COVID-19 Stringency Index [9] and HEV measurements at the Munich Wastewater Treatment Plant (WWTP) Gut Großlappen, indicating that HEV concentrations there tended to be lower when lockdown measures were stricter. In contrast, no statistically significant relationship with lockdown status was evident either in reported case data or at an upstream sampling station with a smaller catchment area. The WWTP findings suggest that HEV transmission may be more sensitive to social and behavioural changes than previously assumed, but this interpretation remains uncertain given the lack of corroboration across datasets. Thus, the question of whether the observed reductions in wastewater HEV concentrations at Gut Großlappen are consistent with a primarily foodborne infection, or whether they suggest additional routes of transmission, merits further investigation.

Agent-based models (ABMs) offer a rigorous framework to gain insight into this question [10]. ABMs can capture stochasticity at the individual level, making them particularly well-suited to studying the transmission dynamics of HEV, since shedding variability, the rarity of symptomatic cases, and substantial measurement noise all contribute to heterogeneous patterns in wastewater-based as well as clinical surveillance data. Previous modelling efforts for HEV in developed settings are sparse, are generally either cohort- or compartment-based, and involve limited or no parameter inference from data [11, 12, 13]. To our knowledge, no prior modelling studies leverage WBS data, which have only recently become available for HEV [14, 15].

In this study, we develop a network-based ABM of HEV dynamics in Munich, Germany that captures individual-level transmission via consumption of raw or undercooked pork, disease progression, differential case ascertainment by symptom status, viral shedding into the sewer system, and measurement noise. We calibrate our model using Approximate Bayesian Computation (ABC) under three scenarios: fitting to longitudinal wastewater data collected from two sampling stations with different catchment sizes during and after the COVID-19-related lockdowns of 2020–2022; fitting to Munich reported case data over the same period despite substantial under-reporting; and jointly fitting to both datasets to assess whether the mismatch between wastewater and reported-case signals can be explained. This framework allows us to explore whether the reductions in HEV concentrations observed at the WWTP during lockdown periods can be explained solely by changes in foodborne exposure, without invoking person-to-person transmission. It also enables investigation into why such reductions manifest at the larger but not the smaller catchment scale, and under what conditions an analogous decline might become observable in the reported case data. Collectively, our work demonstrates the value of combining wastewater epidemiology with individual-level mathematical modelling to deepen our understanding of pathogens that are difficult to monitor through traditional case-based methods.

## 2 Methods

### 2.1 Data

We utilised a unique longitudinal wastewater surveillance dataset from two sampling locations in Munich, Germany. The Gut Großlappen WWTP, which serves 961,207 inhabitants, provided 24-hour composite samples from November 2021 through November 2023. The Schmidbartlanger site, which serves a 66,914-inhabitant subset of the Gut Großlappen catchment, contributed morning grab samples from July 2020 through November 2023. All wastewater viral concentration measurements were normalised using pepper mild mottle virus (PMMoV) as a human faecal indicator to account for environmental confounders such as rainwater infiltration [16]. Sampling, analysis, and normalisation procedures were described in a previous publication [8]. The only pig slaughterhouse within Munich permanently ceased operations on 30 June 2023 [17]. We consider it unlikely that slaughterhouse waste appreciably influenced WWTP viral loads beforehand, because disposal of animal by-products into municipal sewers is prohibited [18]. Consistent with this, mean normalised HEV concentrations at Gut Großlappen were similar in the first versus last six months of 2023 (4.0 × 10^4^ copies/L vs. 4.5 × 10^4^ copies/L; Welch’s t-test: p = 0.70; Mann-Whitney U-test: p = 0.75). We therefore do not explicitly account for slaughterhouse-related confounding.

Throughout our analyses, we define “lockdown periods” as times when a state of emergency was declared in Bavaria due to the COVID-19 pandemic: 16 March 2020–16 June 2020, 9 December 2020–6 June 2021, and 11 November 2021–11 May 2022 [19]. Thus, the Gut Großlappen dataset spans the third of three lockdown periods, while the Schmidbartlanger dataset spans the second and third. As previously reported [8], the mean normalised HEV concentration at Gut Großlappen during lockdown periods was 1.8 × 10^4^ copies/L, significantly lower than the non-lockdown mean of 4.0 × 10^4^ copies/L (Welch’s t-test: p = 0.01; Mann-Whitney U-test: p = 0.002). In contrast, the mean concentration at Schmidbartlanger during lockdown periods was 2.2 × 10^4^ copies/L, comparable to the non-lockdown mean of 2.5 × 10^4^ copies/L (Welch’s t-test: p = 0.74; Mann-Whitney U-test: p = 1.00). The Schmidbartlanger site also exhibited more variable observed concentrations than Gut Großlappen (Fig. 1a), likely reflecting the disproportionate influence of single shedding events in the smaller catchment.

**Figure 1:**
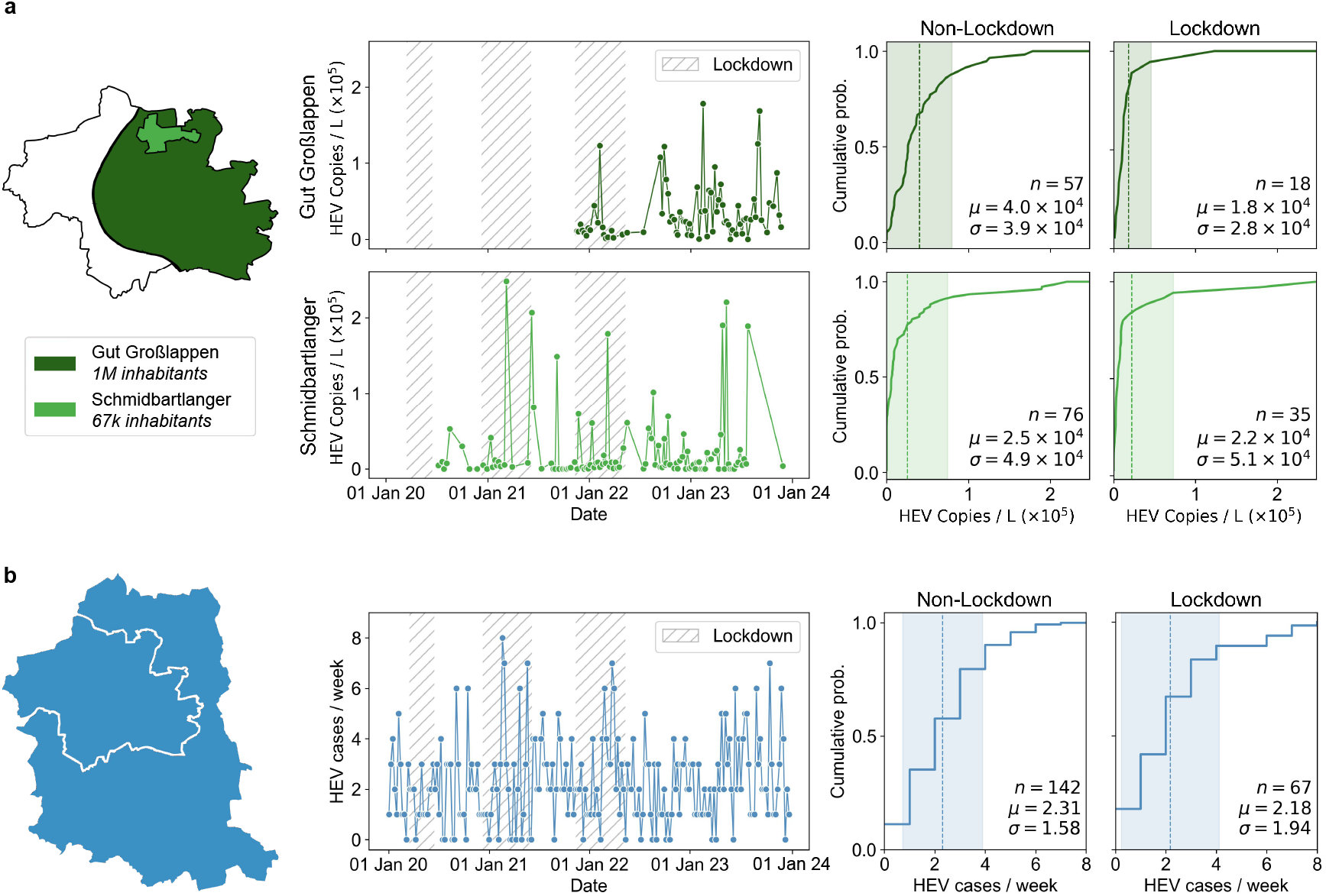
Key data features. **a)** Map of the catchments of the Gut Großlappen WWTP and the smaller Schmidbartlanger sampling site (left) with their longitudinal PMMoV-normalised HEV RNA concentrations (centre). Shaded intervals denote COVID-19 lockdown periods. The corresponding cumulative distributions stratified by lockdown status (right) indicate that concentrations measured at Gut Großlappen were significantly lower during lockdown periods, while those measured at Schmidbartlanger were not noticeably different. Dashed lines denote the mean *µ* and shaded regions show the range spanning one standard deviation *σ* above and below it. **b)** Map of the City of Munich and Munich County (left) alongside the weekly reported HEV case counts for this region, irrespective of clinical status, with lockdown intervals shaded (centre). The corresponding empirical cumulative distributions (right) show no significant difference in weekly case counts between lockdown versus non-lockdown periods.

Weekly reported HEV cases for Munich for 2020–2023 were obtained from an online database of the Robert Koch Institute, Berlin [6], which aggregates data on all notifiable diseases. We considered all reported HEV cases from the City of Munich and Munich County, regardless of clinical status, rather than restricting analyses to cases meeting the national reference definition. Reported incidence did not differ meaningfully by lockdown status (2.18 vs. 2.31 cases per week on average; Welch’s t-test: p = 0.63; Mann-Whitney U-test: p = 0.24) (Fig. 1b).

Together, these descriptive comparisons highlight a divergence across data modalities: the lockdown-associated signal detectable in wastewater at Gut Großlappen is not mirrored at Schmidbartlanger or in the reported case data.

### 2.2 Model overview

To investigate HEV transmission pathways and their modulation by COVID-19-related lockdowns, we developed an ABM comprising one million agents representing the population of the Gut Großlappen WWTP catchment area. The model time frame consisted of 1,461 daily time steps corresponding to 1 January 2020–31 December 2023, covering all three lockdown periods defined in Section 2.1.

Based on the results of a 2021 German dietary survey [20], 90% of agents were designated as omnivores. The remaining 10% were designated as vegan, vegetarian, or pescatarian, and thus not susceptible to foodborne HEV infection. Agents were assigned initial ages according to Munich’s age pyramid as of 2023 and allocated to households following an algorithm replicating five household composition statistics for Munich [21], thereby ensuring realistic household size distributions. Specifically, (i) the average household had 1.8 members; (ii) 17.2% of households contained at least one child; (iii) every household contained at least one adult; (iv) 15.2% of households with children contained only one adult; and (v) 54.9% of households were one-person households. A subset of households was then sampled to represent the Schmidbartlanger catchment area such that its total modelled population equalled 67,000, and viral shedding into wastewater was aggregated at the catchment level (Fig. 2, top).

**Figure 2:**
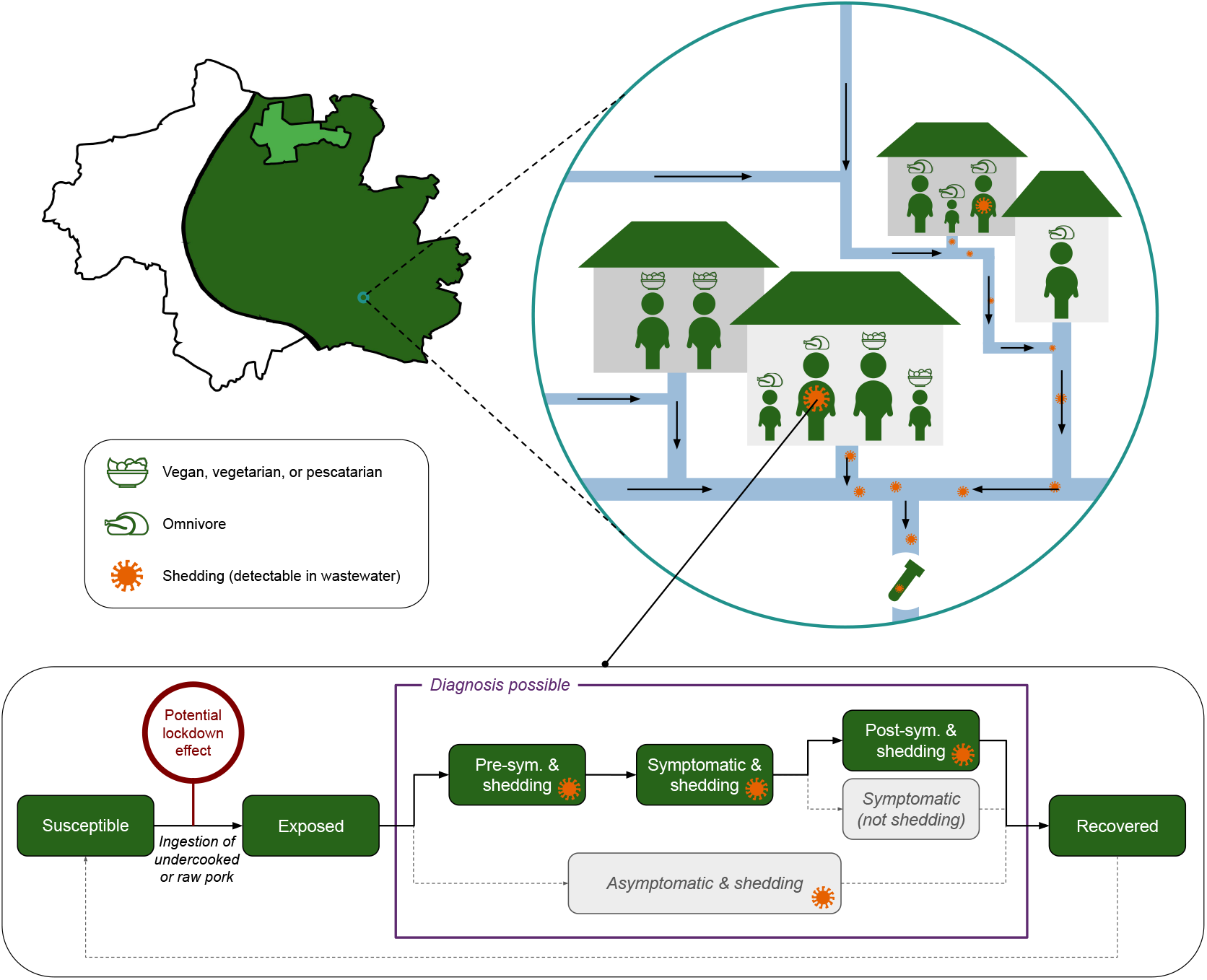
Overview of model structure. The modelled population comprises individuals residing in the catchment area upstream of the Munich WWTP Gut Großlappen (dark green). A subset of these individuals also falls within the smaller catchment area upstream of the Schmidbartlanger sampling station (light green). Agents are designated as vegetarians/vegans/pescatarians (salad icons) or omnivores (poultry icons), with only the latter susceptible to foodborne HEV infection via raw or undercooked pork consumption; the probability of exposure can be modulated by lockdown status. Infected agents progress through the SEIRS-like framework shown in the lower panel, with all states associated with viral shedding into the sewer system indicated by an orange virus icon.

### 2.3 Modelling of transmission and disease progression

Susceptible omnivore agents could be infected with a baseline (non-lockdown) per-day probability of 0.00002. This value was chosen via a one-dimensional calibration sweep to match the estimated pre-COVID-19 annual seroconversion rate in Germany, 5.18 per 1000 person-years [5], by minimizing the absolute difference between simulated and empirical rates under time-invariant and homogeneous risk among omnivores. During lockdowns, this baseline probability was multiplied by an inferred damping factor *l ≥* 0 to capture the effects of any behavioural changes influencing foodborne transmission; the calibrated baseline thus fixes the reference level, making *l* identifiable. The transmission probabilities for all potential person-to-person transmission pathways (within households and via transient social contacts) were fixed to zero to reflect the current understanding of HEV as near-exclusively foodborne in Germany.

The natural history of HEV was represented using a variation on the Susceptible-Exposed-Infectious-Recovered-Susceptible (SEIRS) framework (Fig. 2, bottom), where “Exposed” denotes agents who are infected but not yet shedding. 16.8% of the modelled population started in the recovered, seropositive (immune) state [5], with the remainder starting as seronegative and (if omnivorous) susceptible. Upon infection, agents were stochastically assigned to either an asymptomatic (99%) or symptomatic (1%) clinical pathway [7]. On average, viral shedding began three weeks post-infection and lasted for five weeks [3]. Among symptomatic cases, symptoms typically appeared 40 days after infection [22] and persisted for an average of three weeks [23]. Symptom onset occurred during the shedding period but could conclude before or after shedding ceased. Transitions between disease states were modelled using daily transition probabilities, yielding geometrically distributed durations [24]. Recovered agents could stochastically undergo seroreversion with an annual probability of 0.019 [5] and re-enter the susceptible state, where they could be reinfected under the same parameters as before. Over four simulated years with constant transmission, the seropositive proportion exhibited minimal drift, ranging from 16.8% to 17.6% within a typical simulation.

Explicitly modelling individual faecal shedding trajectories based on serial HEV RNA measurements in stool and/or serum samples [25, 26] did not materially change results; for the sake of parsimony, we therefore modelled only the number of individuals shedding and captured residual variability via a noise model (see Section 2.4).

### 2.4 Modelling of surveillance processes

Diagnosis probabilities were derived from Munich case-to-infection ratios. Based on national HEV sero-conversion estimates [5] and Munich’s 2022 population [27], approximately 9,480 new infections occurred annually during our model period, comprising 95 symptomatic and 9,385 asymptomatic cases. For each calibration scenario involving case reports (see Section 2.5), diagnosis probabilities were computed using the average annual number of reported cases within a selected set of years. Specifically, the average annual number of cases meeting both clinical and laboratory diagnostic criteria in the City of Munich and Munich County was divided by 95 to give the symptomatic diagnosis probability, while the average annual number of cases meeting only the laboratory criteria was divided by 9,385 to give the asymptomatic diagnosis probability.

For each simulated infection, a binomial draw using these diagnosis probabilities determined whether the infection was detected, with the baseline probabilities optionally multiplied by an inferred lockdown diagnosis modifier *r ≥* 0 during lockdown intervals to reflect changes in healthcare-seeking or testing (*r <* 1 reduced detection, *r* = 1 left it unchanged, and *r >* 1 increased it). Report dates were sampled uniformly between shedding onset and recovery or shedding cessation, whichever was later. Simulated reported cases were aggregated by calendar week for comparisons with empirical data.

For each simulated day *t*, the modelled number of persons *n*_*i*_(*t*) shedding into a given catchment area *i ∈ {S, G}* (*S* denoting Schmidbartlanger and *G* Gut Großlappen) with population *N*_*i*_ was mapped deterministically to a preliminary noise-free wastewater concentration *C*_*i*_(*t*) via an inferred catchment-specific scaling factor *s*_*i*_ *≥* 0:

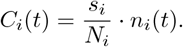

The scaling factor *s*_*i*_ aggregates the effects of the amount of viral material shed per infected person per day and hydraulic dilution by the catchment’s wastewater volume. Random fluctuations in concentration (e.g. due to isolated high-shedding events near the sampler), measurement error, and undetectable observations were represented by a multiplicative noise model with zero inflation. Specifically, undetectable measurements were generated by a Bernoulli indicator *Z*_*i*_(*t*) *∼* Bernoulli(*p*_*i*_(*t*)) with:

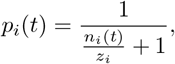

so that the chance of non-detection decayed hyperbolically as the number of shedding individuals increased. The inferred catchment-specific noise parameter *𝓏*_*i*_ *≥* 0 controls the strength of the zero inflation, with smaller values implying fewer non-detections for a given number of shedders. Conditional on a detectable measurement (*Z*_*i*_(*t*) = 0), multiplicative log-normal noise was applied to produce the observed concentration 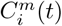:

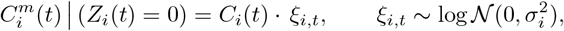

where *σ*_*i*_ *≥* 0 denotes the inferred catchment-specific standard deviation of the log-normal multiplicative error. This error model was chosen over additive Gaussian noise because wastewater concentrations typically exhibit over-dispersion, with measurement variability increasing as concentrations increase [28]. The log-normal formulation captures this mean-variance relationship while preserving positivity. Overall, 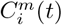 is given by:

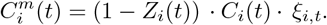

### 2.5 Model calibration

We utilised three contrasting calibration scenarios to explore how well our model could reproduce the wastewater and reported case data streams, both separately and together. The *wastewater-only* scenario solely compared simulated to empirical wastewater concentrations while deliberately disregarding clinical surveillance. The *case-only* scenario, motivated by a counterfactual scenario in which wastewater surveillance data are unavailable, considered only simulated versus empirical reported cases. The *joint* calibration scenario incorporated both datasets. All three strategies utilised the same computational framework, but each used a different distance function and set of free parameters.

In the *wastewater-only* scenario, seven model parameters not directly observable from data were estimated from the wastewater data using Approximate Bayesian Computation with sequential Monte Carlo sampling (ABC-SMC) [29], implemented via the Python package pyABC [30] and using a population size of 200 particles per generation. These parameters were: (i) the lockdown damping factor *l*; (ii) the scaling factors *s*_*S*_ and *s*_*G*_; (iii) the multiplicative noise parameters *σ*_*S*_ and *σ*_*G*_; and (iv) the zero-inflation noise parameters *𝓏*_*S*_ and *𝓏*_*G*_. Broad, uniformly distributed priors were used. Calibration minimised a sum of Kolmogorov-Smirnov (K-S) distances,

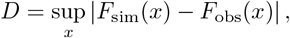

where *F*_sim_ and *F*_obs_ denote the cumulative distribution functions of the simulated and observed concentration values, respectively. One such distance was computed for each combination of sampling location and lockdown status, and these four distances were summed without weighting (treating the shorter lockdown period as equally important). To ensure comparability under irregular sampling, *F*_sim_ was constructed by evaluating simulations only on each catchment’s empirical observation days. Calibration was halted once the composite K-S distance plateaued, the ABC-SMC acceptance rate decreased below 2%, and the 95% credible interval (CI) for *l* ceased to appreciably narrow across generations.

In the *case-only* scenario, the lockdown damping factor *l* was calibrated solely to reported case data using the same ABC-SMC procedure. For this scenario, the lockdown diagnosis modifier *r* was fixed to 1 (i.e. assuming no detection change during lockdowns) for identifiability. The overall diagnosis probabilities were computed as described in Section 2.4 using 2020–2023 averages (51 cases per year meeting both clinical and laboratory diagnostic criteria; 67.5 cases per year meeting only laboratory criteria [6]), yielding diagnosis probabilities of 53.8% for symptomatic and 0.7% for asymptomatic infections. Because the available case counts represent all of the City of Munich and Munich County (population 1.83 million), while the model only simulated the one-million-person Gut Großlappen catchment, these data were first “thinned” to the model scale using binomial subsampling with probability *p* = 1*/*1.83. We generated 200 alternative observation sets to ensure stability, representing plausible realizations of reported catchment incidence. For each lockdown status, the K-S distances between the simulated weekly case distribution and each thinned dataset were computed and then averaged, yielding one mean K-S distance per status. The sum of the lockdown and non-lockdown mean distances served as the objective function for calibration.

Finally, in the *joint* calibration, the model was fit to both wastewater and reported case data simultaneously. The same ABC-SMC framework was used again, and all seven free parameters from the *wastewater-only* calibration were estimated alongside the lockdown diagnosis modifier *r*. For this scenario, we recalculated the diagnosis probabilities for non-lockdown periods as described in Section 2.4 using only 2022–2023 averages, reflecting more stable post-pandemic conditions (56.5 cases per year meeting clinical and laboratory diagnostic criteria; 68 cases per year meeting only laboratory criteria [6]). This resulted in very similar diagnosis probabilities for both symptomatic (59.6%) and asymptomatic infections (0.7%). The composite distance function was extended to include four wastewater-related K-S distances (by lockdown status and sampling location) and two case-related K-S distances (by lockdown status), for a total of six additive terms.

All parameter estimation runs were performed on a machine equipped with dual AMD EPYC CPUs (2× EPYC 7F72 at 3.20 GHz / EPYC 7443 at 2.85 GHz). In the following, we report the mode and the 95% credible intervals (CIs) for the estimated parameters. We used the percentile-based credible interval calculation method implemented in pyABC.

## 3 Results

### 3.1 Calibration to wastewater supports reduced transmission during lockdowns

To quantify the lockdown-associated change in HEV transmission implied by our wastewater data and to test whether reduced foodborne exposure alone can explain it, we first analysed the results of our *wastewater-only* calibration scenario. We monitored convergence of the ABC-SMC algorithm across generations (Fig. 3a) and terminated the calibration after generation 19 in accordance with our stopping criteria; although the acceptance rate fell below 2% at generation 17, we ran two additional generations to allow the 95% CI for *l* to stabilise after a transient widening. Across all generations, approximately 109,000 simulations were executed, requiring a total of about 225 wall-clock hours on 48 cores.

**Figure 3:**
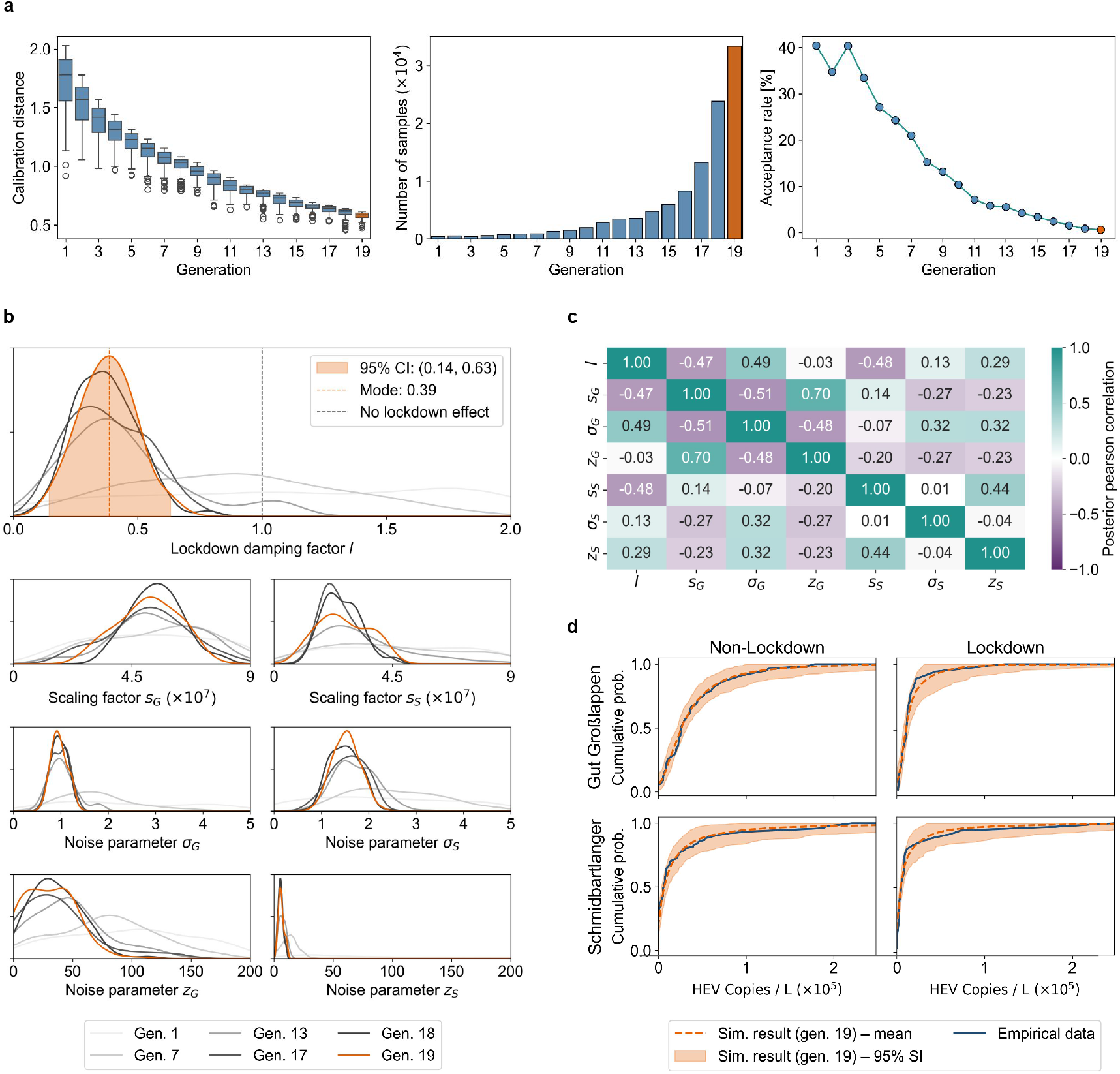
Results of model fitting to wastewater data. **a)** Calibration diagnostics across generations: distribution of distances between simulated and empirical wastewater concentrations by lockdown status and sampling location; total number of samples; and the corresponding acceptance rate. The final generation is in orange. **b)** Posterior distributions of the seven calibrated parameters across selected generations. The x-axes reflect the bounds of the (uniform) prior distributions. For the lockdown damping factor *l*, the shaded orange region denotes the 95% CI of the final-generation posterior, with the inferred posterior mode indicated by a dashed orange line. **c)** Pearson correlation coefficients among the seven calibrated parameters’ posterior distributions, illustrating inter-dependencies among the scaling, noise, and zero-inflation components for each sampling location. **d)** Agreement between simulated and empirical cumulative distributions of PMMoV-normalised HEV concentrations by sampling location and lockdown status. The shaded orange area represents the 95% SI across the 200 accepted simulations from the final ABC generation, and the orange line shows the corresponding mean estimate.

Posterior estimates for the lockdown damping factor *l* indicate a clear reduction in foodborne transmission during lockdown periods. The posterior mode is 0.39 and the 95% CI is 0.14–0.63, which does not overlap with 1 (no lockdown effect) (Fig. 3b).

Posterior distributions for the catchment-specific wastewater scaling and noise model parameters are well-identified (Fig. 3b), with modest correlations across parameters within each sampling location (Fig. 3c). Since the Schmidbartlanger concentrations have a higher variance, the maximum a posteriori (MAP) estimate for *σ*_*S*_ (1.54) is higher than that for *σ*_*G*_ (0.92), while the MAP estimate for *s*_*S*_ (2.25×10^7^) is lower than that for *s*_*G*_ (5.21 × 10^7^). Because multiplicative noise is asymmetric, a higher noise level must be offset by a lower scaling factor to avoid an upward bias in the simulated concentrations.

Simulated cumulative distributions from the final accepted generation closely reproduce the empirical wastewater distributions across both catchments and lockdown categories, with the observed data consistently falling within the 95% simulation intervals (SIs) across the 200 accepted simulations. The simulations capture key empirical features, including the heavy right tails representing occasional high outlier concentrations, the high frequency of low detected concentrations, a clearly visible difference by lockdown status for Gut Großlappen, and the absence of such a difference for Schmidbartlanger (Fig. 3d).

### 3.2 Calibration to case data alone fails to identify a lockdown-associated transmission change

To assess whether lockdown-related differences in HEV transmission can also be inferred in the absence of wastewater data, we analysed the results of the *case-only* calibration scenario. In this case, the ABCSMC algorithm converged more rapidly (Fig. 4a), reflecting the lower dimensionality of this inference task, which involves only a single free parameter. Approximately 32,000 simulations were executed in total, requiring about 65 wall-clock hours.

**Figure 4:**
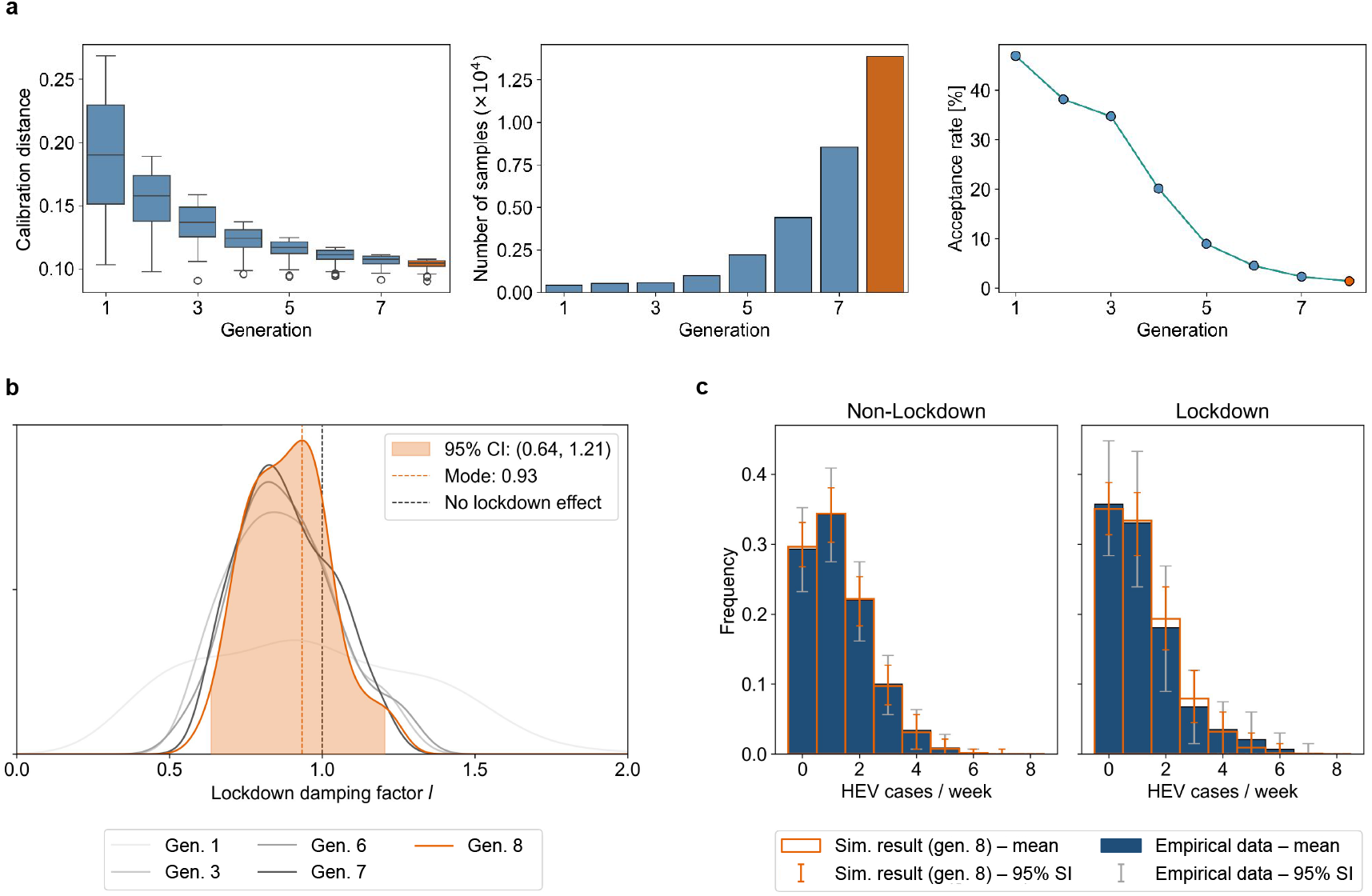
Results of model fitting to reported case data. **a)** Calibration diagnostics across generations: distribution of distances between simulated and empirical weekly reported case counts by lockdown status; total number of samples; and the corresponding acceptance rate. The final generation is in orange. **b)** Posterior distributions of the calibrated lockdown damping factor *l* across selected generations. The shaded orange region denotes the 95% CI of the final-generation posterior, with the inferred posterior mode indicated by a dashed orange line. **c)** Agreement between simulated and empirical distributions of weekly HEV case notifications, stratified by lockdown versus non-lockdown periods. Orange bars represent the mean of the 200 accepted simulations from the final ABC generation, with orange error bars indicating the associated 95% SI. Dark blue bars and black error bars show the mean and 95% SI obtained by thinning the empirical case observations to the modelled population size via binomial subsampling.

For this scenario, the posterior for the lockdown damping factor *l* is broad and covers the no-effect value of 1 (Fig. 4b), which is expected based on the lack of a lockdown-related signal in the reported case data (Fig. 1b). Thus, case data alone do not provide evidence for a reduction in HEV transmission during lockdown periods.

Simulated case distributions from the final generation align well with the right-skewed empirical histograms for both lockdown and non-lockdown conditions, indicating that the model can accurately capture the low-incidence, over-dispersed nature of HEV clinical surveillance data (Fig. 4c).

### 3.3 Joint calibration to wastewater and case data can reconcile discordant empirical signals

To determine whether the discordant lockdown patterns observed across wastewater and case surveillance can be reconciled within a single mechanistic model, we analysed the results of our *joint* calibration scenario. Calibration diagnostics stabilised by generation 17 (Fig. 5a) and calibration was halted at this point, as matching the *wastewater-only* generation count was prohibitively time-consuming. Approximately 44,000 simulations were executed in total, requiring about 90 wall-clock hours.

**Figure 5:**
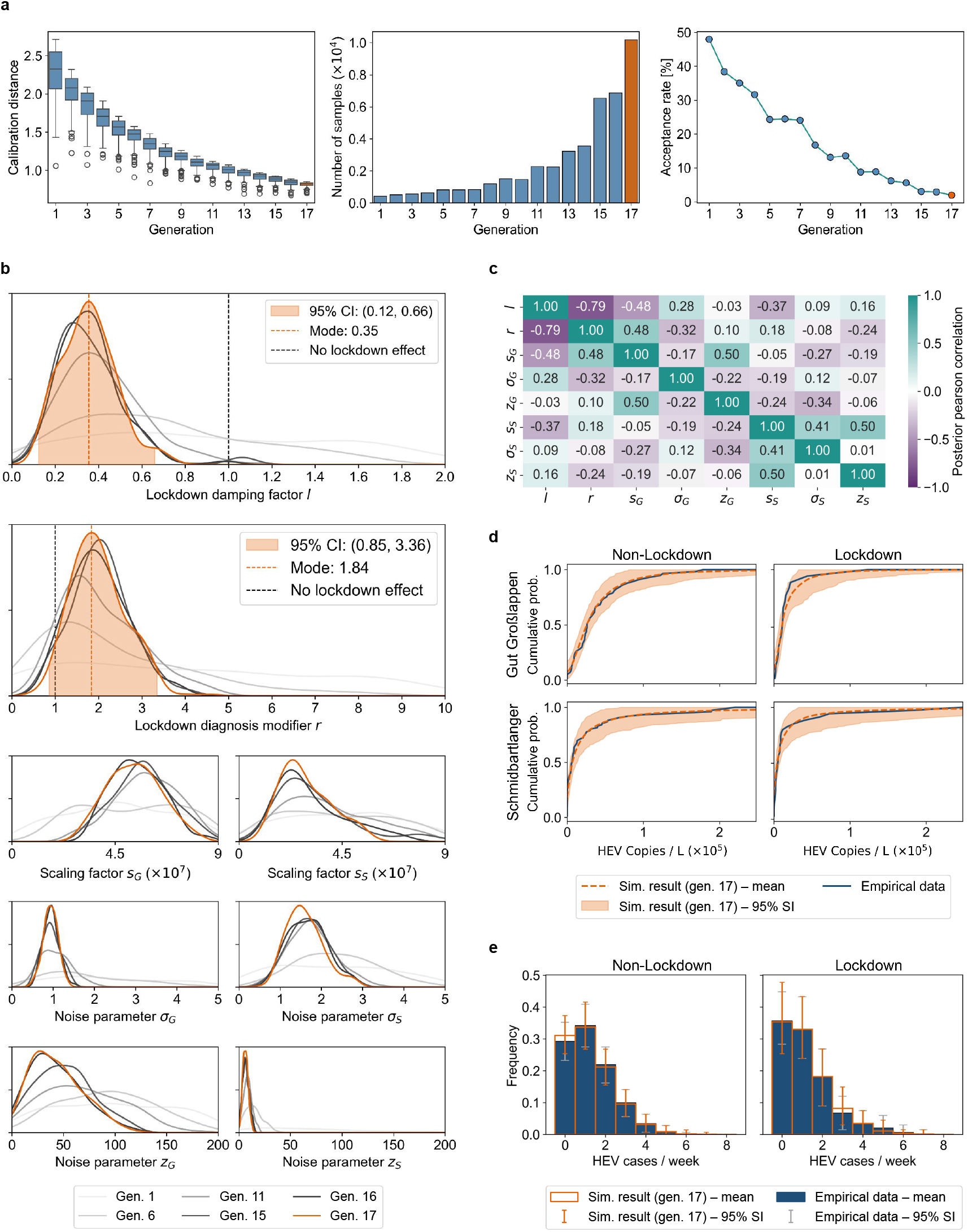
Results of joint model fitting. **a)** Calibration diagnostics across generations. **b)** Posterior distributions across selected generations. **c)** Posterior Pearson correlations among the calibrated parameters. **d)** Agreement between simulated and empirical cumulative distributions of HEV concentrations. The orange line and shading represent the mean and 95% SI across the 200 accepted final-generation simulations. **e)** Agreement between simulated and empirical distributions of weekly HEV cases. Orange bars and error bars show the mean and 95% SI across the 200 accepted final-generation simulations; blue bars and black error bars show the mean and 95% SI of the thinned empirical observations.

Consistent with the *wastewater-only* calibration, the *joint* calibration posterior for the lockdown damping factor *l* indicates a substantial reduction in foodborne transmission during lockdowns. The posterior mode is 0.35 (compared to 0.39 previously), and the 95% CI again excludes the no-effect value of 1. The lockdown diagnosis modifier *r* has a posterior mode of 1.84 (95% CI: 0.85–3.36), suggesting a higher probability of detection during lockdowns, although the 95% CI is wide enough to include 1 and therefore remains compatible with no change (Fig. 5b). Interpreting *l*, a value of 0.35 is compatible with scenarios in which 65% of transmission normally occurs via restaurants and this component drops to zero during lockdowns, or scenarios in which all transmission is restaurant-related but decreases by 65%, as two extreme illustrative examples. Given that the Google COVID-19 Community Mobility Reports for Bavaria document reductions of up to 90% in the “retail and recreation” mobility category (which includes visits to restaurants and cafés) during lockdown periods [31], the magnitude of reduction implied by intermediate scenarios between these two extremes is plausible.

Parameter correlation patterns are generally consistent with those from the *wastewater-only* calibration. Additionally, there is a strong negative correlation between the two lockdown effect parameters *l* and *r* (Pearson correlation coefficient = -0.79, p *<* 0.001), reflecting a trade-off between reduced transmission and increased diagnosis likelihood (Fig. 5c). This interdependency between *l* and *r* illustrates how nonstationary ascertainment can offset true transmission signals and thereby hinder inference from case notifications alone.

The final-generation simulations reproduce empirical wastewater concentration distributions for both catchments and lockdown statuses (Fig. 5d), consistent with the *wastewater-only* calibration, and match the empirical weekly reported case count distributions with high fidelity (Fig. 5e), consistent with the *case-only* calibration. Among the 200 accepted simulations, few show statistically significant lockdown-related differences at the 0.05 significance level in weekly reported cases (15 by Welch’s t-test; 17 by the Mann-Whitney U-test) or in Schmidbartlanger wastewater measurements (43 by Welch’s t-test; 29 by the Mann-Whitney U-test). In contrast, most simulations show significant differences in Gut Großlappen wastewater measurements (133 by Welch’s t-test; 156 by the Mann-Whitney U-test).

Together, these results indicate that the combination of a clear lockdown-related signal in wastewater from the Gut Großlappen catchment and the absence of such a signal in the smaller Schmidbartlanger catchment and in the case surveillance data is consistent with a true underlying reduction in HEV transmission during lockdowns. In the reported case data, this reduction may have been offset by a modest increase in diagnosis probability, although this effect is uncertain; at the smaller catchment scale, it is likely obscured by higher measurement noise.

### 3.4 Detecting lockdown differences in case data would require substantially higher reporting

Because transmission damping (*l <* 1) and heightened diagnosis probabilities (*r >* 1) during lockdowns have opposing effects on reported case counts, they can offset one another, leaving little or no observable lockdown signal in the case data even when transmission truly changes (Section 3.3). This motivates the question of how close the “status quo” non-lockdown diagnosis probabilities must be to the lockdown diagnosis probabilities (treated as a practical upper bound) for the reported case data to consistently exhibit a lockdown-related difference, as observed in the Gut Großlappen wastewater data. We therefore conducted an ascertainment scenario analysis using the most likely parameter values from the *joint* calibration. Specifically, the MAP estimates of the lockdown damping factor and diagnosis modifier, *l*_MAP_ = 0.37 and *r*_MAP_ = 1.87, were obtained using a two-dimensional kernel density estimate of their joint posterior distribution, marginalising out the remaining parameters, and used as fixed inputs. Non-lockdown diagnosis probabilities were then systematically increased toward lockdown-level values using a linear interpolation parameter *α ∈* [0, 1], where *α* denotes the proportion of the full 87% increase required to reach lockdown-level reporting. Thus, *α* = 0 corresponds to the reference non-lockdown diagnosis probabilities (0.7% for asymptomatic and 59.6% for symptomatic infections), while *α* = 1 applies the full multiplier *r*_MAP_ = 1.87 (yielding 1.3% for asymptomatic and 100% for symptomatic infections).

For each value of *α*, 100 stochastic simulations of the HEV model were run, and the resulting per-simulation distributions of weekly reported case counts under lockdown versus non-lockdown conditions were compared using Welch’s two-sample t-tests and Mann-Whitney U-tests. Increasing *α* produced a monotonic, near-linear rise in the mean difference in simulated weekly cases (Fig. 6a, top). For the reference scenario (*α* = 0), almost no simulations yielded a statistically significant lockdown effect at the 0.05 significance level under either statistical test. Significance rates rose sharply between *α* = 0.3 and *α* = 0.8 and plateaued thereafter due to near-saturation (Fig. 6a, bottom). For *α* = 0.8 (corresponding to an increase of about 70% in non-lockdown diagnosis probabilities), 95% of simulations exhibited a significant difference between lockdown and non-lockdown reported cases under both statistical tests. Scenario-specific histograms demonstrate that increasing *α* progressively shifts the non-lockdown weekly case distribution towards higher values, with a clear divergence from the lockdown distribution visible for *α* = 0.7 and *α* = 1.0 (Fig. 6b).

**Figure 6:**
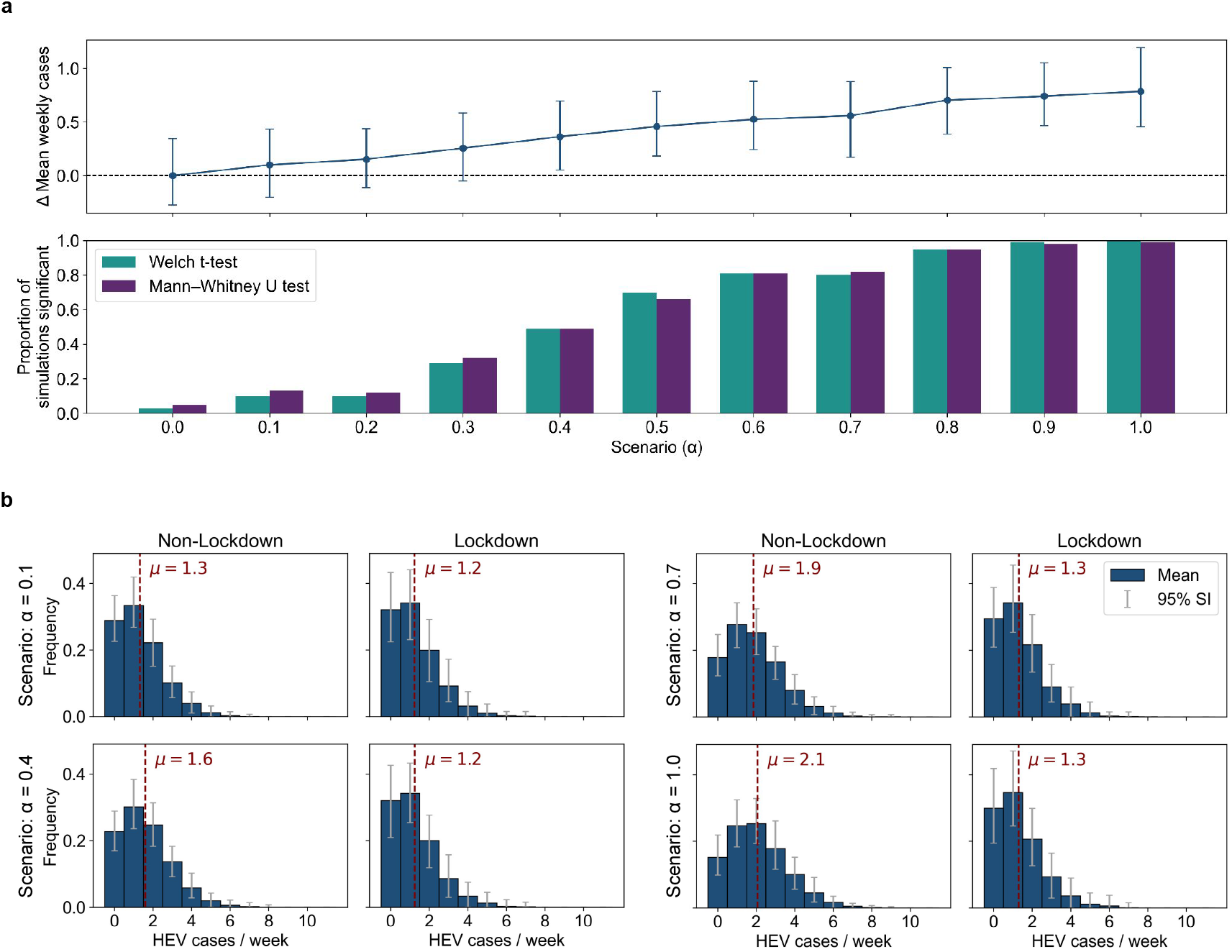
Scenario analysis of non-lockdown diagnosis probability increases. **a)** Impact of progressively increasing non-lockdown diagnosis probabilities towards the lockdown level using the interpolation parameter *α ∈* [0, 1]. The top panel shows the change in mean weekly reported cases under non-lockdown versus lockdown conditions across 100 stochastic simulations for each *α* value, with error bars indicating the 95% SI. The bottom panel shows the corresponding proportion of simulations for which the difference in weekly case counts between lockdown and non-lockdown periods is statistically significant (*p <* 0.05), based on Welch’s two-sample t-tests (teal) and Mann-Whitney U-tests (purple). **b)** Representative distributions of weekly reported HEV case counts by lockdown status for four selected diagnosis scenarios (*α* = 0.1, 0.4, 0.7, 1.0). Bars and error bars indicate the mean and 95% SI respectively across the 100 simulations per scenario. Red dashed lines indicate the average weekly case count *µ*, computed within each simulation and then averaged across simulations.

Taken together, these results indicate that a substantial (*≥* 70%) increase in non-lockdown case ascertainment, particularly among asymptomatic individuals, would be required to generate robust differences in reported case incidence comparable to those observed in the wastewater signal, highlighting the role of surveillance biases in reported case data.

## 4 Discussion

This study demonstrates how integrating WBS data into a flexible agent-based model of HEV dynamics can yield insights that are difficult to obtain from conventional case-based reporting alone. We leverage a unique longitudinal wastewater dataset encompassing two catchments with markedly different population sizes and spanning two out of three COVID-19 lockdown periods, creating a natural experiment to assess how behavioural changes may have unintentionally altered the transmission of faecal-oral pathogens alongside the respiratory disease being targeted. Our results reconcile seemingly contradictory patterns across surveillance modalities, providing evidence for a true reduction in HEV transmission during lockdowns that was statistically clear in the wastewater data from the WWTP Gut Großlappen but obscured at the smaller catchment scale by high measurement variability and in reported case data by the inherent stochasticity of the low-case regime, possibly combined with lockdown-related shifts in care-seeking or testing. Together, our findings highlight both the strengths and limitations of WBS for detecting subtle population-level epidemiological shifts.

The central insight of this work is the added value of wastewater data when under-detection is profound. In our setting, diagnosis probabilities are not only low but may also be non-stationary due to altered healthcare-seeking behaviour and testing practices during the COVID-19 pandemic, resulting in case notifications that may not reliably reflect true transmission dynamics. Indeed, our joint inference suggests that case ascertainment may have increased during lockdown periods, although the lockdown diagnosis modifier is estimated imprecisely and remains compatible with no change. While many health services were disrupted during the pandemic, plausible mechanisms nonetheless exist for increased HEV ascertainment. For example, liver abnormalities were prevalent among COVID-19 patients [32], which may have prompted more frequent viral hepatitis rule-out testing and, by extension, increased incidental detection of HEV. Our scenario analysis indicates that for clinical surveillance to reliably detect a lockdown effect on HEV transmission, non-lockdown diagnosis probabilities would need to increase by about 70%, bringing them very close to the corresponding lockdown values. This would effectively demand pandemic-level testing intensity outside a pandemic context, which is unrealistic. More generally, our use of a binary lockdown diagnosis modifier is a pragmatic simplification; continuous variation in ascertainment would likely be more realistic but unidentifiable from the available data, further complicating the interpretation of case-based surveillance. WBS, in contrast, reveals population-level trends by capturing viral material shed by all infected individuals, regardless of symptom status, care-seeking behaviour, or clinical prioritisation. Economies of scale also make it cost-effective to add pathogens to wastewater surveillance systems already established for SARS-CoV-2 [33, 34]. However, wastewater signals are shaped by interacting biological, behavioural, and infrastructural processes, and are therefore not straightforward to interpret [35, 36]. Mechanistic modelling frameworks like ours can therefore help disentangle true epidemiological changes from sampling artifacts or stochastic variability.

We also demonstrate how strongly the detectability of transmission changes in wastewater depends on the size of the contributing population. The larger Gut Großlappen catchment exhibited a clear lockdown-associated decline in HEV concentrations that was robust to both dichotomous and continuous definitions of lockdown severity, including the COVID-19 Stringency Index [8]. In contrast, the upstream Schmidbartlanger catchment, approximately 15-fold smaller, showed no discernible difference. Our modelling results indicate that this discrepancy can be explained without invoking different underlying transmission dynamics. When fewer individuals contribute to a sewershed, the wastewater signal can become dominated by stochasticity due to individual high-shedding events near the sampling location and to sewage that is less well-mixed. Accordingly, larger catchments provide more temporally stable indicators of transmission, as aggregation averages out individual shedding trajectories as well as daily and weekly behavioural cycles [37, 36]. In the emerging-disease context, smaller catchments may indeed be advantageous for detecting localised outbreaks [38]. But HEV is a continuously circulating pathogen whose RNA signal is notably stable in wastewater [39] (as an example of the persistence of non-enveloped enteric viruses, hepatitis A RNA has an estimated half-life of 25 days in water [40], implying only about 1.4% loss after 12 hours, the upper end of typical sewer travel times in urban Europe [41]). As such, the disadvantages of relatively longer sewer travel times are reduced for viruses like HEV, making large catchments preferable for reliably monitoring perturbations in their transmission.

Finally, our findings add nuance to the prevailing understanding of HEV genotype 3 transmission in developed countries. In these settings, HEV is regarded as a predominantly foodborne pathogen with infections primarily driven by consumption of undercooked pork products, which are widely contaminated [42, 43]. As such, one might expect HEV transmission to be insensitive to lockdown requirements, which are intended to disrupt interpersonal contact patterns rather than food supply chains. However, in calibrating our model without invoking person-to-person transmission, we estimate that foodborne exposure during lockdowns fell to about 35% of the reference level. Plausible mechanisms include shifts from restaurant dining to home cooking [44, 45]; fewer opportunities to purchase high-risk products like raw minced pork sandwiches, which are typically sold as ready-to-eat items in butcheries and bakeries [46, 47]; reduced fresh meat consumption in favour of shelf-stable foods to economise or limit grocery trips [44, 48]; and enhanced hygiene practices during food preparation prompted by heightened infection-control awareness [45]. Our modelling results thus do not contradict existing evidence from serological studies [49], outbreak investigations [50, 51], and risk factor analyses [52, 53] showing that direct human-to-human transmission of HEV genotype 3 is exceedingly rare in Europe [54].

Although our model aims for realism by incorporating variation in shedding and symptom durations as well as diagnosis delays, key aspects of HEV natural history remain poorly characterised, particularly the temporal dynamics and inter-individual variability of viral shedding magnitude. As such, we do not explicitly model differences in shedding rates between individuals, including the presence of high-shedding outliers or variation by infection severity. Our use of a constant per-time-step seroreversion probability could be refined to incorporate an initial period of near-complete protection followed by time-varying waning if more data become available on refractory period duration and subsequent waning speed; however, given low seroreversion rates relative to our model time frame, we would not expect this to materially change our results. In addition, certain sources of heterogeneity in wastewater data are not modelled mechanistically. For example, Schmidbartlanger utilised grab rather than composite sampling, although these strategies have been shown to yield comparable signals for other viruses such as influenza [55]; and as a smaller upstream catchment, Schmidbartlanger may also be more affected by daily and weekly fluctuations in the contributing population than the WWTP. In the present analysis, these complexities are all absorbed into the two noise parameters per site. While this simplification is unlikely to affect our main conclusions, more detailed future modelling of different noise processes could improve the interpretation of site-specific differences.

While ABC has clear advantages as a flexible and widely used likelihood-free calibration framework, it also has practical limitations that could in the future be mitigated by emerging methods [56]: it is computationally intensive, lacks a single definitive convergence diagnostic, and requires hand-crafted discrepancy metrics (here, an unweighted rather than adaptive distance scheme). Independently of these algorithmic considerations, comparisons between the *case-only* and the *joint* calibration scenarios are limited by differences in model settings. The *case-only* calibration reflects a counterfactual situation in which no wastewater data are available, meaning there would be no a priori reason to postulate a lockdown-specific change in both transmission and reporting. Allowing both the lockdown damping factor and diagnosis modifier to vary freely in this setting would introduce substantial identifiability problems; we therefore fixed the latter to one (no effect) in the case-only calibration. As a result, the two sets of calibration results should not be interpreted as independent estimates of the same parameters but rather as complementary scenarios illustrating how inference may change when additional data streams are considered.

Finally, our inference assumes homogeneous changes in foodborne exposure across the omnivore population during lockdowns. Given that contamination of pork products with HEV is widespread, this is a reasonable approximation. However, vegetarianism is associated with younger age and higher education in Germany [57], so university districts in Munich may be at systematically lower risk of HEV; the same is likely true for neighbourhoods with a high prevalence of pork avoidance for religious reasons, even if residents are classified as omnivores for consuming other meats. Conversely, our assumption that vegetarian diets entail zero risk of HEV transmission may be overly strict, since HEV RNA has occasionally been detected in plant-based dishes such as seaweed salad [58]. We also do not explicitly model within-household clustering of infection due to a family consuming the same undercooked pork dish, for example. Future extensions that incorporate demographic and dietary differences at the household and neighbourhood level could better characterise local variations in risk.

In summary, calibrating an agent-based model against wastewater measurements can reveal epidemiological changes in the transmission of HEV, a profoundly under-reported infection, that are not apparent in clinical case data. Our findings provide a foundation for future work to quantify the burden of other under-detected pathogens based on integrated surveillance strategies.

## Data Availability

All data and custom code used in this study is available on GitHub at https://github.com/karina-d-wallrafen/HEV-model-analysis.

## Author contributions

K.W.S.: conceptualization, methodology, software, formal analysis, investigation, writing – original draft, writing – review & editing, visualization; J.J.: data curation, writing – review & editing; N.S.: methodology, writing – review & editing, visualization; M.S.: validation, resources, writing – review & editing; J.J.W.: validation, resources, writing – review & editing, supervision, funding acquisition; A.W.: validation, resources, writing – review & editing, supervision, funding acquisition; J.H.: validation, writing – review & editing, supervision, project administration, funding acquisition.

## Implementation and availability

Our model of HEV dynamics was built in R using the EpiModel software platform [59]. All data and custom code used in this study is available on GitHub at https://github.com/karina-d-wallrafen/HEV-model-analysis.

